# Bridging the gap: Enhancing the generalizability of epigenetic clocks through transfer learning

**DOI:** 10.1101/2025.02.14.25322080

**Authors:** Lan Luo, Lulu Shang, Jaclyn M. Goodrich, Karen E. Peterson, Peter X-K Song

## Abstract

Changes in DNA methylation patterns exhibit a high correlation with chronological age. Epigenetic clocks, developed through statistical models that estimate epigenetic age using the methylation levels of cytosine-guanine dinucleotide (CpG) sites, have emerged as powerful tools in understanding aging and age-related diseases. Despite their popularity, the generalizability of these clocks across diverse populations remains a challenge. We find that some of the widely used epigenetic clocks, such as Horvath’s clock (Horvath, 2013) and PedBE clock (McEwen et al., 2020) do not perform well in our target cohort. This lack of representativeness raises concerns about applying these clocks to quantify biological age in distinct demographic and ethnic groups. In addition, the feature space between existing clocks and our target data is different: most existing clocks are trained with data from older platforms, such as the Illumina HumanMethylation450 BeadChip (450K). In contrast, our target data are profiled with a more recent Illumina HumanMethylationEPIC BeadChip (EPIC) array. To address these gaps, we propose a transfer learning framework to adapt existing epigenetic clocks to underrepresented populations, using shared knowledge from diverse datasets. Furthermore, we develop imputation- and DNN-based methods for feature adaptation between existing clocks and our target data. Using data collected from 593 blood samples from a cohort of children and adolescents in the ELEMENT study, we find that our proposed transfer learning methods greatly improve the prediction performance compared to applying existing clocks directly. Performance is further enhanced by using the CpG sites profiled on the EPIC array. Our methodology showcases the potential to bridge the gap between different DNAm datasets and different profiling platforms, thus improving the applicability of epigenetic clocks in diverse population groups and contributing to more accurate aging research.

## 1 Background

Changes in the methylation state of cytosine-guanine dinucleotide (CpG) sites have been shown to be highly correlated with chronological age. Thus, DNA methylation (DNAm) patterns are being increasingly used to gain insight into aging and aging-related diseases. Epigenetic clocks are statistical models used to estimate epigenetic age based on DNA methylation at specific CpG sites.

In the rapidly evolving field of epigenomics, various epigenetic clocks have been developed with diverse sources of training data. Since the initial model by Bocklandt et al. (2011), this field has evolved towards more robust and sophisticated models. One of the most popular epigenetic clocks was developed by Horvath (2013), termed Horvath’s clock, which uses DNAm at 353 CpGs selected by elastic net penalized regression model (Zou and Hastie, 2005) to predict chronological age. Later, several other clocks have been developed, including Horvath’s Skin and Blood clock (DNAm AgeSkinBlood) (Horvath et al., 2018) and Hannum’s clock (Hannum et al., 2013). These clocks are referred to as the *first-generation clock* for predicting chronological age. To better capture biological age and health-related outcomes, the *second-generation clock* has been developed to predict the risk of age-related diseases and lifespan and assess the biological impacts of lifestyle and environmental factors. Examples include Levine’s PhenoAge clock to predict lifespan and susceptibility to diseases (Levine et al., 2018), GrimAge clock to predict mortality risk and risks related to smoking behavior (Lu et al., 2019), as well as the DunedinPACE clock to quantify the pace of aging (Belsky et al., 2020). The field continues to evolve, with the implementation of new predictive models and the development of species- and tissue-specific epigenetic clocks.

One major limitation of existing epigenetic clocks is the lack of generalizability. An epigenetic clock developed to estimate age or age-related diseases in a population may not translate into other populations with divergent demographic and socioeconomic profiles (Tyrrell et al., 2021; Taylor et al., 2018; Munafó et al., 2018). We find that some of the existing epigenetic clocks, including the popular Horvath clock, and two other clocks developed for children (Pediatric-Buccal-Epigenetic (PedBE) clock (McEwen et al., 2020) and Wu’s clock (Wu et al., 2019)), are not really suitable for our target study, which is a cohort of adolescents aged 8-18 years; see the correlation coefficient in Table 1. Such incompatibility raises a serious question about the validity of existing aging clocks in quantifying biological age and age acceleration among *say*, children, midlife women, or patients with specific diseases like diabetes and Alzheimer’s disease.

**Table 1:** Summary of some of the existing clocks and their performances in our target dataset. “CpGs in clock” represents the number of CpGs in each of the listed clocks, “missing CpGs (%)” indicates the number and percentage of CpGs that are not profiled in the target dataset, and “correlation” shows the correlation coefficient between the predicted and the true chronological ages when applying each of these clocks to the target dataset.

| Clock | CpGs in clock | Missing CpGs (%) | Correlation ($R$) | Reference |
| --- | --- | --- | --- | --- |
| Horvath | 353 | 25 (7.1%) | 0.55 | Horvath (2013) |
| Hannum | 71 | 11 (15.5%) | 0.27 | Hannum et al. (2013) |
| Levine | 513 | 9 (1.8%) | 0.26 | Levine et al. (2018) |
| SkinHorvath | 391 | 9 (2.3%) | 0.55 | Horvath et al. (2018) |
| PedBE | 94 | 1 (1.1%) | 0.44 | McEwen et al. (2020) |
| Wu | 111 | 4 (3.6%) | 0.62 | Wu et al. (2019) |
| TL | 140 | 20 (14.3%) | 0.13 | Lu et al. (2019) |
| BLUP | 319607 | 22803 (7.1%) | 0.56 | Zhang et al. (2019) |
| EN | 514 | 9 (1.8%) | 0.54 | Zhang et al. (2019) |

Ideally, datasets used for training, validation, and subsequent analysis should be as similar as possible in the sense that they should all have been generated with the same technology, under the same laboratory conditions, and represent the same underlying population (Teschendorff and Horvath, 2025). In such an ideal setting, the results of applying an epigenetic clock should be directly interpretable as chronological age or biological age. Therefore, if the predicted value differs remarkably from the true value, the clock is not suitable for the target data. Mounting evidence has shown that the generalizability of models trained in a particular population can be low due to a substantial amount of heterogeneity in underlying populations (West et al., 2017; Landry et al., 2018; Kraft et al., 2018; Duncan et al., 2019). At least in terms of racialized groups, two recent reviews have reported that epigenetic data are predominantly from individuals of European heritage (Watkins et al., 2023). Our preliminary results also show that both the PedBE clock and the Wu clock, trained primarily with individuals of Caucasian descent, perform less well in our target cohort collected from Mexico City (Perng et al., 2019). These inconsistencies may be due in part to some clocks including loci known to be differentially methylated by country of birth and racialized group. Applying an existing epigenetic clock to a population with different demographic characteristics and environmental exposures may lead to inappropriate inferences, especially when this target cohort is poorly represented in the training data used to derive the clock (Taylor et al., 2018; Munafó et al., 2018; Watkins et al., 2023).

To advance biological age prediction, it is crucial to improve the performance of existing epigenetic clocks in underrepresented populations so as not to distort scientific inference. One may use study-specific data only to build a prediction model. However, the sample size in a target study can be limited, especially when data are collected from underrepresented populations. This is more of a concern for high-dimensional DNAm data, especially when deep neural networks (DNNs) are used to construct epigenetic clocks (Levy et al., 2020; Galkin et al., 2021; de Lima Camillo et al., 2022). To account for such heterogeneity and lack of representation, we propose to use transfer learning to leverage the shared knowledge learned from diverse populations or bulk tissue samples to a specific target cohort, maybe an underrepresented population or a specific tissue or cell type, so that comparable model performance can be achieved with much less data for training. The concept of transfer learning provides an organic way to integrate pre-existing models into the target prediction task. The basic idea is to use existing epigenetic clocks trained with abundant source data to improve age prediction performance in the target cohort, so a completely new clock does not have to be developed from scratch with possibly limited target data. This is extremely important when dealing with high-dimensional DNAm data where the number of predictors far exceeds the sample size, and even more so when DNNs are used to capture complex interactions among CpG sites (de Lima Camillo et al., 2022).

As a motivating example, we will use the DNA methylation dataset of children and adolescents in the ELEMENT cohort (Perng et al., 2019) to illustrate our proposed framework to transfer existing clocks to a target study. In addition to the heterogeneity in the two underlying populations between the source and target datasets, these two data sources also differ in terms of their profiling platforms. Training data for building most existing clocks are profiled with 27K/450K platforms, but our ELEMENT methylation data are obtained by the more recent EPIC array covering over 850,000 CpGs distributed genome-wide. In other words, the number of CpG sites profiled by the EPIC array is more than twice of the existing clocks. Therefore, in addition to the difference in the selected CpGs and their weights, the dimension of the input feature vector also differs. To the best of our knowledge, the differences in the feature dimensions between the source and target domains, especially when dealing with DNAm datasets, have not been addressed before.

To bridge the gaps between existing clocks and a target study, we propose a transfer learning framework in epigenetic clocks to address heterogeneity in both the underlying populations and profiling platforms. Our proposed framework has the following advantages over applying an existing clock directly: (a) it leverages rich information extracted by existing clocks from massive and diverse DNAm data; (b) it allows both the addition or exclusion of CpG sites that are no longer predictable in the target cohort; (c) it bridges the gap between an existing clock and the target cohort so that the transferred clocks have greatly improved prediction performance in the target dataset. We describe the generic methods for transferring existing epigenetic clocks in Section 2. The application of the proposed methods to the ELEMENT dataset is included in Section 3. Finally, we include a discussion, outline the limitations, and suggest areas of future research in Section 4.

## 2 Methods

Our goal is to take advantage of the epigenetic clocks developed on a certain data source to increase its predictive performance on the target cohort. One of the key benefits of transfer learning is that it can reduce the amount of data required to train an effective model. This is particularly advantageous in training epigenetic clocks because DNAm datasets typically contain ultra-highdimensional features with a relatively small number of subjects. The idea is to transfer the selected CpGs and their weights in existing clocks to a new target dataset, and further fine-tune the model with the target dataset.

### 2.1 Formulation of epigenetic clocks

In this paper, we focus on the first-generation clock, which is a high-dimensional linear regression model. Formally, an existing clock can be written as

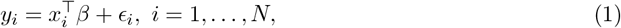

where 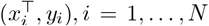 are independent samples, *β* ∈ ℝ^*p*^ is the coefficient or weight vector of interest, and *ϵ*_*i*_, *i* = 1, …, *N* are independently distributed random noises with 𝔼 [*ϵ*_*i*_ | *x*_*i*_] = 0. In the first-generation clock, *y*_*i*_ ∈ ℝ is typically the chronological age of the *i*th subject, and *x*_*i*_ ∈ 𝒳^*S*^ ⊂ ℝ^*p*^ is a *p*-dimensional vector containing the methylation levels at different CpG sites with 𝒳^*S*^ denoting the feature space of the source domain. In a high-dimensional setting, *p* can be much larger than *N*. For example, depending on the profiling platforms, the dimension *p* can be hundreds of thousands, while *N* is mostly on the scale of several hundreds. However, most CpGs are not strongly correlated with age and *β* is often assumed to be sparse such that the number of nonzero elements of *β*, denoted by *s*, is much smaller than *p*. We summarize some of the well-known existing epigenetic clocks and the number of selected CpG sites in the first two columns in Table 1.

Regardless the differences in both the number and the loci of the selected CpGs in existing clocks, they are mostly trained via the linear regression model with elastic-net penalty (Zou and Hastie, 2005). The objective function is given by

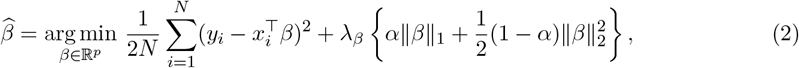

where *α* ∈ [0, 1] is a mixing parameter that balances the weight of lasso (*α* = 1) and ridge (*α* = 0) penalties, *λ*_*β*_ ≥ 0 controls the overall strength of the penalty. The true regression coefficient is assumed to be 𝓁_0_-sparse, which satisfies ∥*β*∥_0_ = *s* ≪ *p*. In general, among hundreds of thousands of CpG sites being profiled, only hundreds of them contribute to the chronological age prediction.

### 2.2 Gaps between existing clocks and a target dataset

Our goal is to improve the prediction performance of existing epigenetic clocks in a target study cohort, for example, our motivating dataset from the ELEMENT study. In the field of epigenetic clocks, the weights or coefficient estimates 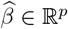 in existing clocks are typically accessible. However, due to data heterogeneity arising from different demographic features, cell types, profiling platforms, and pre-processing techniques, existing clocks trained from a different source dataset may perform poorly in the target cohort, see the correlation coefficient in Table 1.

Alternatively, we may train a completely new clock from scratch using our target dataset only. Assume the target dataset consists of independent samples 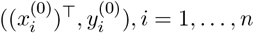, a new clock trained with target data only can be formulated as 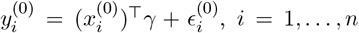, where 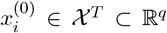. But this strategy loss valuable information from existing clocks trained with possibly a much larger dataset assembled from a diverse population.

In epigenetic clocks, we expect at least some of the predictive CpGs to be shared across different study populations, tissues, or cell types. Therefore, it would be reasonable and beneficial if we could borrow information from the existing clock to optimize the prediction performance in the target dataset. Recently, transfer learning methods have been developed for high-dimensional regression models (Li et al., 2022; Tian and Feng, 2023; Gu et al., 2024). However, existing work typically assumes the same feature space between the source and target domain, i.e. 𝒳_*S*_ = 𝒳_*T*_ ∈ ℝ^*p*^ and *p* = *q*, but the conditional distribution of *y* given *x* differs. Transfer learning of this type is referred to as *homogeneous transfer learning*. In the case of high-dimensional linear models, their respective coefficients *β* and *γ* differs. To characterize such a discrepancy, they typically introduce a bias-correction term *δ* = *γ* − *β* which can be estimated through the following objective function:

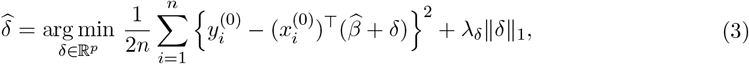

where ∥*δ*∥_1_ is a penalty term depending on the 𝓁_1_-norm distance between *β* and *γ*, and *λ*_*δ*_ is a tuning parameter that can be selected via cross-validation. With such a penalization, the weight vector 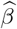 extracted from an existing clock is used to guide the learning of the target parameter *γ*. After obtaining 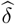, the updated coefficient 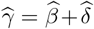 adapts the weights learned from the source data to the target data. However, such a transfer learning framework is built under the assumption that 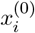 and *x*_*i*_ are of the same dimension *p*. Due to the development of profiling arrays, newer datasets typically contain twice the CpG sites compared to older platforms. Most existing work, when combining datasets profiled on 27K, 450K and EPIC arrays, uses only the overlapped or shared CpG sites across different platforms (McEwen et al., 2020; de Lima Camillo et al., 2022). Such a naive way of bridging the gap in feature dimensions between source and target domain apparently will loss rich and valuable information provided by the newer and higher-resolution profiling EPIC array. To make full use of the methylation information extracted from the high-resolution platforms, we plan to retain the entire vector 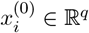 rather than taking a subset of overlapped CpGs, and the transfer learning task in our setting with 𝒳_*S*_ 𝒳_*T*_ is termed as *heterogeneous transfer learning*.

### 2.3 Feature alignment via imputation

To highlight the difference in feature spaces between 𝒳_*S*_ and 𝒳_*T*_, we use a different notation *z*_*i*_ ∈ 𝒳_*T*_ ⊂ ℝ^*q*^ to denote the vector containing methylation levels at CpGs profiled on newer EPIC array. We remark that *q > p* because the newer EPIC array has expanded coverage and includes more methylation sites than the older 450K or 27K arrays. To mitigate the discrepancy in feature dimensions, we introduce a new mapping *h* : *z* → *x*. The function *h*(*z*) can be viewed as a transformation from the target domain to the source domain to align their feature spaces.

A natural way is to impute the methylation levels at the CpG sites used in an existing clock via k-nearest neighbors. The methylation levels of neighboring cytosines tend to be spatially correlated, implying that the nearby CpG sites often exhibit similar methylation patterns (Eckhardt et al., 2006). According to Figure 1 (upper panel), which shows a snapshot of the aligned CpGs by EPIC array and those profiled by 27K/450K arrays, we can see that the loci indicated by the blue bars (EPIC array) are mostly close neighbors to the red bars (27K/450K arrays). Therefore, given the methylation profiles obtained from the EPIC array, we can hypothetically impute the methylation levels at CpG sites as if they were profiled by the lower-resolution 450K or 27K array, which is beneficial for feature alignment between the source and the target domains.

**Figure 1:**
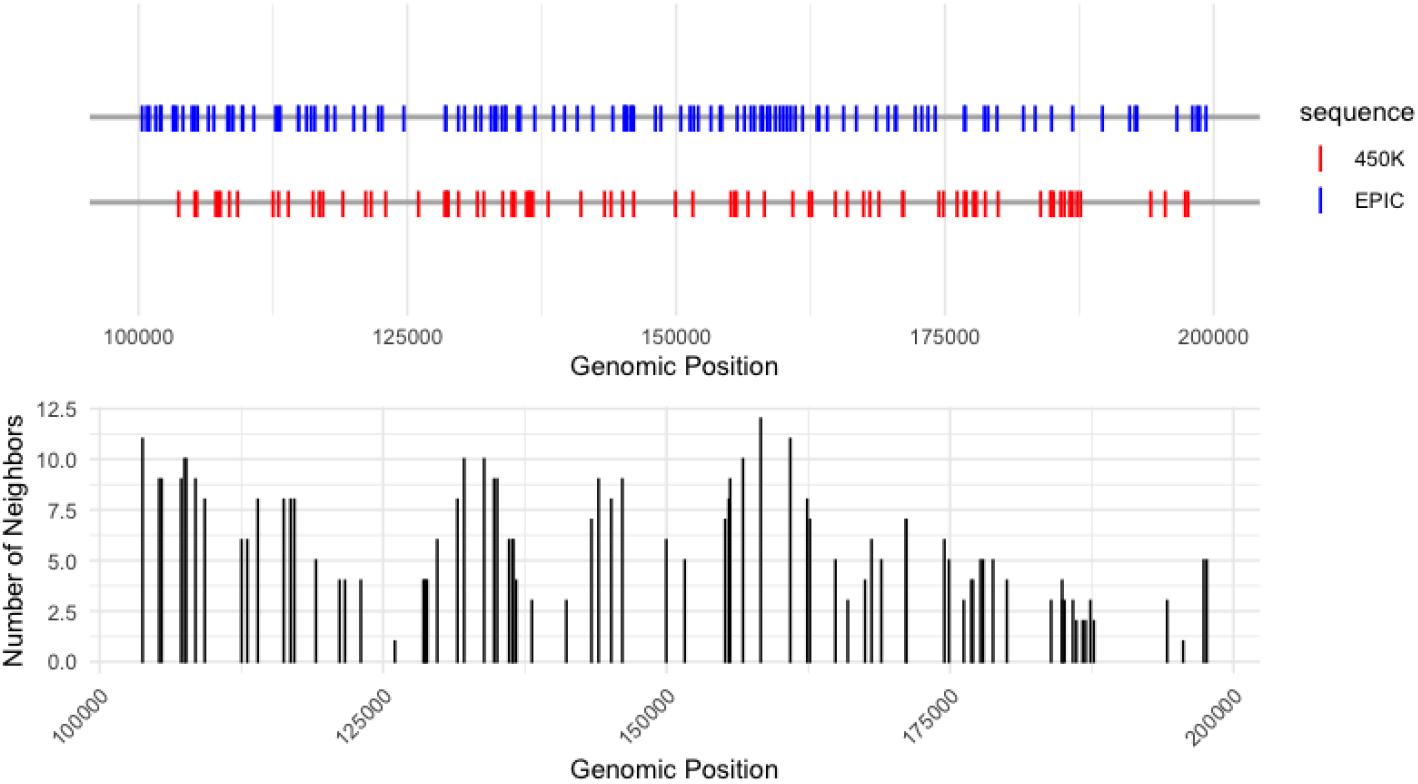
Upper panel: CpG sites in ELEMENT data profiled with EPIC array (blue) v.s. CpG sites by 450K/27K array (red); lower panel: number of neighbors from EPIC array for imputing the methylation levels at each of the CpG sites in 450K/27K platform.

The upper panel in Figure 1 also shows that the CpG sites are not uniformly distributed across the genome, and the density of CpG sites can vary significantly between different genomic regions. To account for these characteristics, we choose to set a distance threshold *d* rather than choose the number of neighbors ad hoc. In this case, the number of neighbors that are used is adaptively determined based on the local density of the CpG sites. The threshold *d* is chosen in a data-driven way so that most CpG sites in 450K or 27K arrays have at least one neighbor. Given a fixed *d*, a diagram of the number of neighbors used for each CpG site is shown in Figure 1 (lower panel).

Let 𝒞_450K_ be the index sets of unique identifiers of CpG sites in the source datasets profiled on a 450K array, and 𝒞_EPIC_ denote the set of CpGs in the target data profiled on an EPIC array, where the number of CpGs in 𝒞_450K_ and 𝒞_EPIC_ are *p* and *q*, respectively. Our goal is to find an explicit form of *h*(*·*) that takes *z* ∈ ℝ^*q*^ as input and output *x* ∈ ℝ^*p*^. For *k* = 1, …, *p*, we will iterate the following steps to complete the transformation from *z* to *x*. First, we extract the genomic position of the *k*th element in *x*, denoted by *j*_*k*_ ∈ 𝒞_450K_. In a special case where the position *j*_*k*_ is also profiled by the EPIC array, we set the methylation level *x*_*k*_ = *z*_*k*_*′* if *j*_*k*_*′* = *j*_*k*_ where *j*_*k*_*′* ∈ 𝒞_EPIC_ is the genomic position of the *k*′th element in *z*. Otherwise, we impute it with neighboring information. Given a distance threshold *d*, we find the neighbors for imputation by considering a set of positions in 𝒞_EPIC_ that are within a distance of *d* from the position *j*_*k*_. More specifically, for every *k*^*′*^ = 1, …, *q*, we calculate its distance to the position *j*_*k*_, defined by *d*(*j*_*k*_, *j*_*k*_*′*) = |*j*_*k*_*′* − *j*_*k*_|, and then find *k*^*′*^ such that *d*(*j*_*k*_, *j*_*k*_*′*) ≤ *d*. Next, we define a weight variable *w*_*k*_*′* such that *w*_*k*_*′ >* 0 if *j*_*k*_*′* ∈ [*j*_*k*_ − *d, j*_*k*_ + *d*] and *w*_*k*_*′* = 0 otherwise for *k* = 1, …, *q*. Lastly, *x* = [*x*_1_, …, *x*_*p*_]^⊤^ = *h*(*z*) = [*h*_1_(*z*), …, *h*_*p*_(*z*)]^⊤^ ∈ ℝ^*p*×1^ where 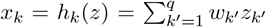 and 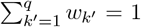. Here, the positive weights are pre-defined, which can lead to a simple average or a weighted average where the closer neighbors contribute more heavily. Note that we drop the subject index when extracting the genomic position, because subjects from the same dataset, either source or target datasets, are typically profiled on the same platform and the probe indexes do not vary by subject.

Then the bias-correction term *δ* is obtained by the following objective function:

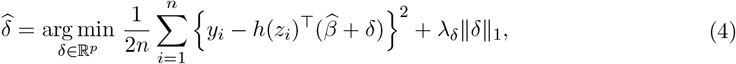

where *h*(*z*_*i*_) ∈ ℝ^*p*×1^ is the vector of methylation levels as if our target data were profiled b y the lower-resolution 450K or 27K array.

### 2.4 Feature adaptation via deep neural networks

Alternatively, the mapping from *z* to *x* can be learned in a data-driven way through DNNs. Our goal is to take the levels of methylation at different C pG s ites (denoted *z* ∈ ℝ^*q*^) i n t he EPIC matrix as input to predict chronological age, with the unknown mapping function denoted by *f* (·) : [0, 1]^*q*^ → ℝ. The basic unit of a neural network is a neuron, which takes input values, processes them using weights and biases (intercepts), and produces an output value. A multilayer neural network or a DNN consists of multiple layers of interconnected neurons with a specific network architecture (*L, d*) where the number of layers *L* is called the depth of the network, while the number of neurons per layer *d*_𝓁_ is the width of the network. The input layer is denoted by 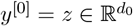 with *d*_0_ = *q*, and the weights from layer 𝓁 to 𝓁 + 1 is denoted by 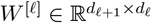, the bias vector is 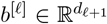 for 𝓁 = 0, …, *L* − 1. To simplify the notation, we extend the activation function: *σ* : ℝ → ℝ to vector-valued inputs, by applying it component-wise such that *σ* : ℝ^*d*^ → ℝ^*d*^. Then the feature vector of the layer 𝓁 in a DNN can be represented by

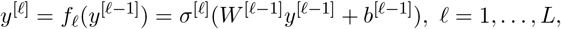

where the activation function *σ*^[𝓁]^ may differ from layer to layer. The output of the DNN is therefore *y*^[*L*]^ = *f* (*z*) = *f*_*L*_ ° *f*_*L*−1_ ° … ° *f*_1_(*z*).

However, such a generic architecture is equivalent to training a DNN from scratch and it loses valuable information as well as interpretability in existing epigenetic clocks. To take advantage of the knowledge learned from existing epigenetic clocks, we propose fixing t he w eights o f t he layer just before the last, with index (*L* − 1), referred to as the “penultimate layer”. Specifically, we set the number of neurons in the penultimate layer as *d*_*L*−1_ = *s* and the weights in the last layer as 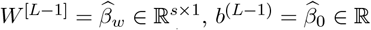 where 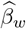 is the weight vector corresponding to the selected CpG sites *s* and 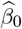 is the intercept in an existing clock. Moreover, the activation function in the last layer is linear.

The newly proposed DNN architecture is shown in Figure 2. Given the difference in resolution between the source and target platforms, the initial layers of our model extract features from the higher-resolution data and project them to the penultimate layer, which might already be effective in making predictions based on knowledge learned from the lower-resolution data. It is worth noting that by fixing the weights of the penultimate layer, we are able to transfer prior knowledge learned from existing epigenetic clocks. The loss function in this case can be expressed as

**Figure 2:**
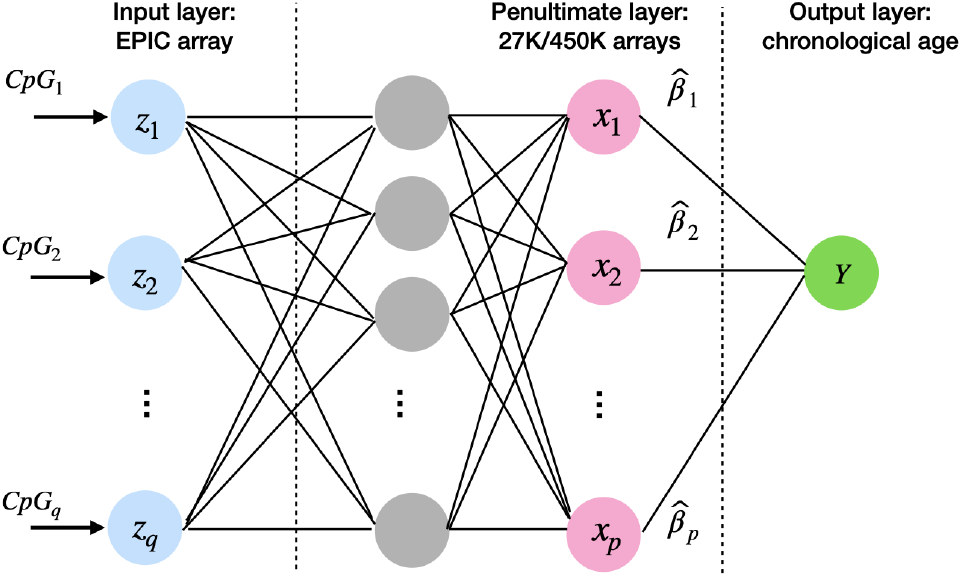
Diagram of the DNN structure for feature adaptation. The input takes methylation levels at CpG sites profiled on the EPIC array. It is then processed through several hidden layers, with the penultimate layer being a learned low-dimensional representation that is equivalent to the selected CpGs in an existing clock.

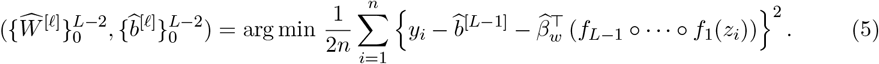

During training, we search for parameter values 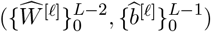 that minimize loss and therefore map the input *z* profiled on EPIC array to chronological ages *y* as closely as possible. In particular, the penultimate layer, represented by *y*^[*L*−1]^, acts as a higher-level representation of the source feature space 𝒳_*S*_, and the weighted combination of the units in this layer through *β*_*w*_ is expected to predict the chronological age. This penultimate layer serves as a bridge between the input from the high-resolution EPIC array and the desired output of chronological age. More importantly, it enables knowledge transfer from existing clocks. Additionally, we note that training a DNN to directly learn the transformation function from EPIC array to 450K or 27K array is infeasible because most training datasets are profiled with either higher- or lower-resolution platforms, not both. Therefore, setting lower-resolution feature representations as the penultimate layer is the key to bridging the gap between existing clocks and the input from the higher-resolution EPIC array.

## 3 Data Application

### 3.1 ELEMENT data collection and preprocessing

Our motivating dataset is from the Early Life Exposures in Mexico to ENvironmental Toxicants (ELEMENT) study (Perng et al., 2019). ELEMENT is a longstanding study that originally recruited mother-child pairs into three sequential cohorts between 1994 and 2003 in Mexico City. The original goals of ELEMENT were to study the impact of prenatal lead exposure on developmental outcomes. Existing epigenetic clocks have been used as biomarkers of health and links between exposure and outcomes in these children (Jansen et al., 2021; Ehlinger et al., 2023; Halabicky et al., 2024). The epigenetic data in ELEMENT come primarily from blood leukocyte samples collected during late childhood and adolescence, with ages ranging from 8 to 18 years.

DNAm was profiled in 610 samples using the Infinium EPIC platform. The raw image files were read with the R package `minfi` (Fortin et al., 2017), and quality control and normalization were performed using the R package `ENmix` (Xu et al., 2017) for data from all batches. The probes were removed if at least 5% of the samples were not detected (p-value *>* 10^−16^ compared to the background; 32,752 probes removed). Background correction was performed with noob and dye bias correction with RELIC followed by quantile normalization (Pidsley et al., 2016). Probes that are known to be cross-reactive (using the list from Pidsley et al. (2016); 43,256 probes removed), have SNPs in the CpG or single-base extension site (based on probe annotation information from the manufacturer) were removed for a final CpG site count of 762,657. After removing missing values in age, a total of 593 subjects with 762,657 CpGs were profiled in our target dataset.

### 3.2 Applying different methylation-based clocks to ELEMENT data

As a baseline comparison, we first apply an existing transfer learning method by restricting ourselves to shared CpG sites between our ELEMENT data and some of the popular clocks. The clocks we include in the comparison are (i) Horvath’s clock (Horvath, 2013), (ii) PedBE clock (McEwen et al., 2020), and (iii) Wu’s clock (Wu et al., 2019). The first clock is one of the most widely used for adults, and the latter two clocks are developed for children and adolescents. They should be suitable starting points given that our ELEMENT data is also extracted from a cohort of children and adolescents.

For performance evaluation, we consider the two commonly used metrics: (a) correlation coefficient (*R*) between predicted (*ŷ*) and true chronological age (*y*), and (b) median absolute error (MAE) between *y* and *y*. Since transfer learning relies on splitting the entire dataset into training and test sets, we apply the aforementioned three clocks to the test set in this section as a fair baseline evaluation.

We first split the ELEMENT dataset into training and test sets with a commonly used ratio: 80% for training and 20% for test sets. First, as a reference, we train a completely new clock from scratch with the target data (ELEMENT data) only; the resulting correlation coefficient is 0.80, and the MAE is 0.85. The penalty parameter selected by 10-fold cross-validation is *λ*_*β*_ = 0.011, and the number of selected CpG sites is 360. We remark that our expectation is a bit different from conventional homogeneous transfer learning in that the transferred clocks will significantly outperform a clock trained with target data only. The reason is mainly due to the difference in feature space: when training a new clock with target data only, we use the information extracted from the EPIC array directly, which includes probes that are more predictable of chronological age than those probes used by the older platforms (Pidsley et al., 2016). Therefore, by transferring those existing clocks trained on lower-resolution platforms, we hope to achieve a comparable or slightly better performance than that achieved by training a new clock using target data only.

For applying existing clocks, since we extract the learned weights directly, we do not train any new model with the training set, and the performances are evaluated with the test set. The resulting correlation coefficient and MAE are shown in the “Original” column in Table 2. The correlation coefficients between the predicted and the true ages are around 0.5 for all three clocks, which are much lower than those in their original datasets used for training these clocks. For example, in Horvath (2013), the correlation coefficient reported in its test set is 0.96, and it is 0.98 in both McEwen et al. (2020) and Wu et al. (2019). Such a huge discrepancy sheds light on the heterogeneity in datasets collected from different cohorts, and applying an existing epigenetic clock arbitrarily to a target study cohort with different environmental exposures and social characteristics can lead to inappropriate interpretation. This observation motivates us to consider the transfer of knowledge learned from existing clocks to a target study cohort.

**Table 2:**
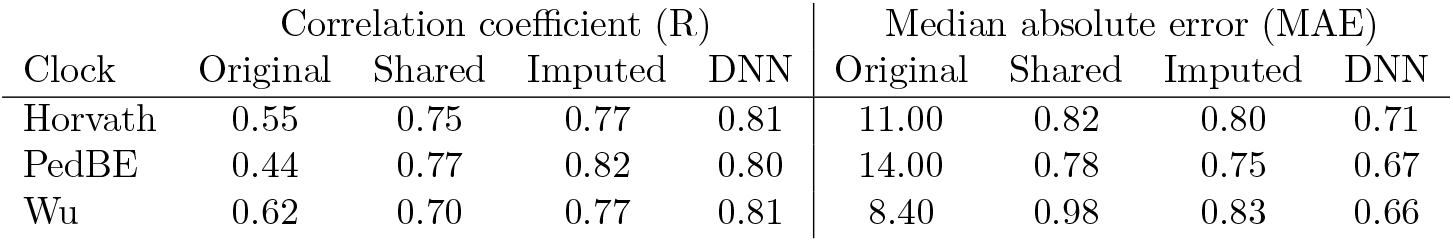
Performances of different methods in terms of correlation coefficient and median absolute error between the predicted outcome and the true chronological age. (i) Original: direct application of existing clocks; (ii) Shared: transfer learning across shared CpGs only; (iii) Imputed: transfer learning after feature alignment via imputation; (iv) DNN: transfer learning via DNN.

| Clock | Correlation coefficient (R) |  |  |  | Median absolute error (MAE) |  |  |  |
| --- | --- | --- | --- | --- | --- | --- | --- | --- |
|  | Original | Shared | Imputed | DNN | Original | Shared | Imputed | DNN |
| Horvath | 0.55 | 0.75 | 0.77 | 0.81 | 11.00 | 0.82 | 0.80 | 0.71 |
| PedBE | 0.44 | 0.77 | 0.82 | 0.80 | 14.00 | 0.78 | 0.75 | 0.67 |
| Wu | 0.62 | 0.70 | 0.77 | 0.81 | 8.40 | 0.98 | 0.83 | 0.66 |

### 3.3 Transfer learning with shared CpGs

The set of CpG sites profiled by the EPIC array in the ELEMENT data may not contain some of the CpG sites in existing clocks, although the EPIC array profiles more CpGs than the 27K or 450K arrays. To perform a transfer learning task and update the weights from an existing clock, we may start by considering their overlapped CpGs. As summarized in Table 1, very few CpGs in those three clocks are not profiled in the ELEMENT data, so we may not lose much information by restricting ourselves to their shared subsets.

It is worth noting that most existing clocks only share their nonzero CpG sites and the corresponding estimated coefficients. To perform transfer learning in a high-dimensional setting, we also have to recover those unselected CpG sites, as they might be predictive in our target dataset. A total of 21, 368 probes that have been used to train the Horvath clock are publicly available in (Horvath, 2013). For the PedBE clock and Wu’s clock, we download their training data from the GEO database and follow the preprocessing steps used in their papers; the resulting numbers of probes are narrowed down to 22, 164 and 22, 233, respectively. After taking an intersection with the probes in our target data profiled in the EPIC array, the input feature dimension becomes *p* = 19, 589, *p* = 16, 387, and *p* = 20, 166 for these three clocks, respectively. Note that the intercept is included in the values of *p*.

This homogeneous transfer learning task is achieved by estimating the bias correction term *δ* using the objective function of equation (3). We solve this optimization problem with the `glmnet` package in `R` where the tuning parameter *λ*_*δ*_ is chosen as the one that minimizes the 10-fold crossvalidation error with the `cv.glmnet()` function. Then the updated weight vector is 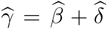, and it is applied to the samples in the test set for performance evaluation. For each of the clocks being transferred, we summarize their respective *λ*_*δ*_ values and number of selected CpGs below:

(i) Horvath’s clock: *λ*_*δ*_ = 0.015, and the number of selected CpG sites is 527; (ii) PedBE clock: *λ*_*δ*_ = 0.019, and the number of selected CpGs of the transferred clock is 262; (iii) Wu’s clock: *λ*_*δ*_ = 0.028, and the number of selected CpGs is 230. Venn diagrams showing the intersection between the CpG sites selected by the original clock and those selected by the transferred clock is shown in Figure 3.

**Figure 3:**
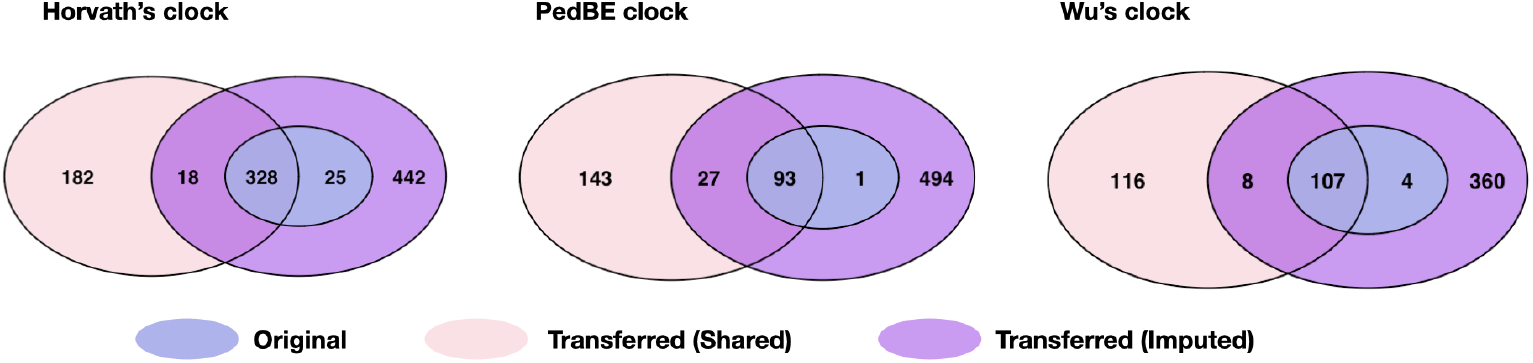
Venn diagrams of the selected CpG sites by (a) the original clock, (b) the transferred clock based on shared CpGs and (c) the transferred clock with imputed CpGs.

The transferred clocks versus their original versions are plotted in Figure 4. The correspondences between DNAm age and chronological age are shown in blue dots for the transferred clocks and black dots for the original clocks. We notice that even if the transferring is conducted across the shared CpGs, the prediction performances improve a lot: the blue dots are mostly centered around the red dashed line, compared to those black dots. The column “Shared” in Table 2 summarizes the results in these two metrics: (i) the correlation coefficient improves by at least 12.9%, and (ii) the MAE decreases to around 1/10th of its original value for all three clocks.

**Figure 4:**
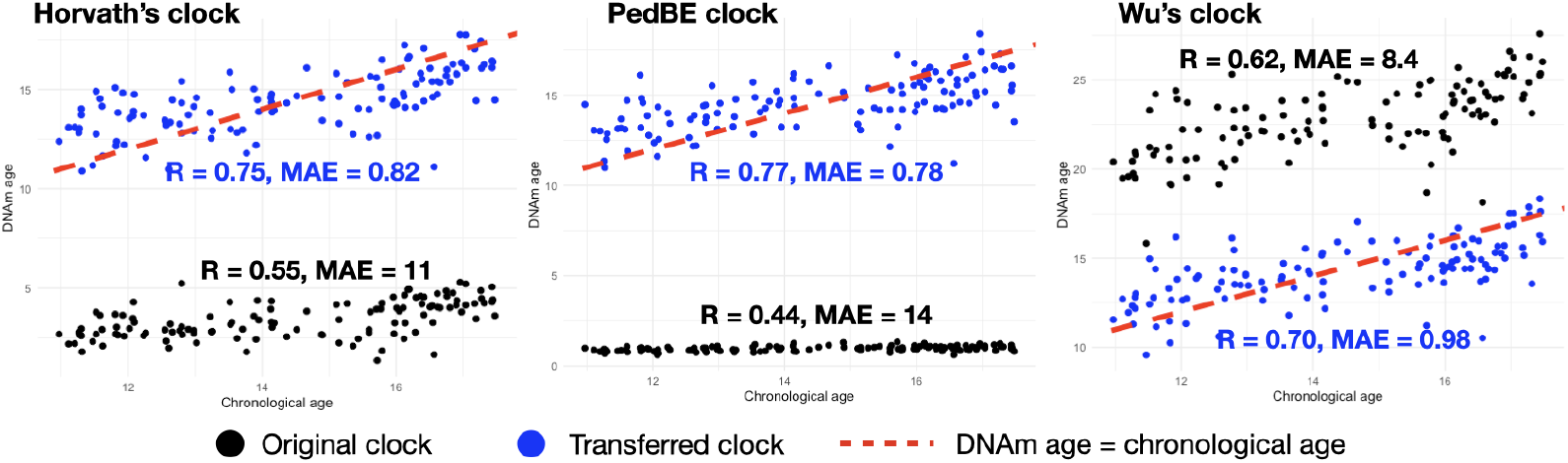
Plots for samples in test sets using the original (black dots) versus the transferred clocks with shared CpGs (blue dots). The 45-degree line (red dashed line) represents perfect agreement between the DNAm age and the chronological age.

### 3.4 Transfer learning after imputation

Now we evaluate the performance of the proposed imputation method in Section 2.3. We apply a distance-based method to determine the number of neighbors to be used, as shown in Figure 1. In this setup, the number of CpGs in the EPIC array is *q* = 762, 657. The numbers of features (including the intercept) we plan to use after imputation are *p* = 21, 369 for Horvath’s clock, *p* = 22, 165 for PedBE clock, and *p* = 22, 234 for Wu’s clock. Instead of choosing a fixed number of neighbors ad hoc, we would like to determine it dynamically by analyzing the distribution of CpG site distances. In the first step, for each CpG site in the 450K or 27K array, we calculate its distances to each of the CpGs profiled by the EPIC array. Then we choose a threshold distance *d* = 2.5 × 10^3^ that determines the number of neighbors that are used for imputation. This distance is chosen such that there is at least one neighbor for the imputation, and the number of neighbors that are used is shown in the lower panel of Figure 1. In the final step, we take an average of the methylation levels in those selected neighboring sites to impute the methylation level at each CpG site as if our target data were profiled by the 450K or 27K array.

The resulting imputed covariate matrix will serve as an input for subsequent transfer learning. In essence, feature alignment is achieved by transforming features from the target domain to the source domain, and the heterogeneous transfer learning task is then converted to a homogeneous transfer learning task. In transferring the Horvath’s clock after feature alignment, *λ*_*δ*_ = 0.0035, and the number of selected CpG sites is 812. In transferring the PedBE clock, *λ*_*δ*_ = 0.0021, and the number of selected CpGs of the transferred clock is 614. For Wu’s clock, *λ*_*δ*_ = 0.0065 and the number of selected CpG sites is 478.

Compared to transfer learning with shared CpG sites, using the PedBE clock as an example, the correlation coefficient after feature alignment is further improved by 9.2%, and MAE is also reduced by 12.6%. See performance in Figure 5 and Table 2. It should be noted that improving prediction performance does not require a large target dataset or accessing the data sets to train the existing clocks. Only CpG unique identifiers used in training existing clocks and methylation levels for CpGs in the target dataset are required.

**Figure 5:**
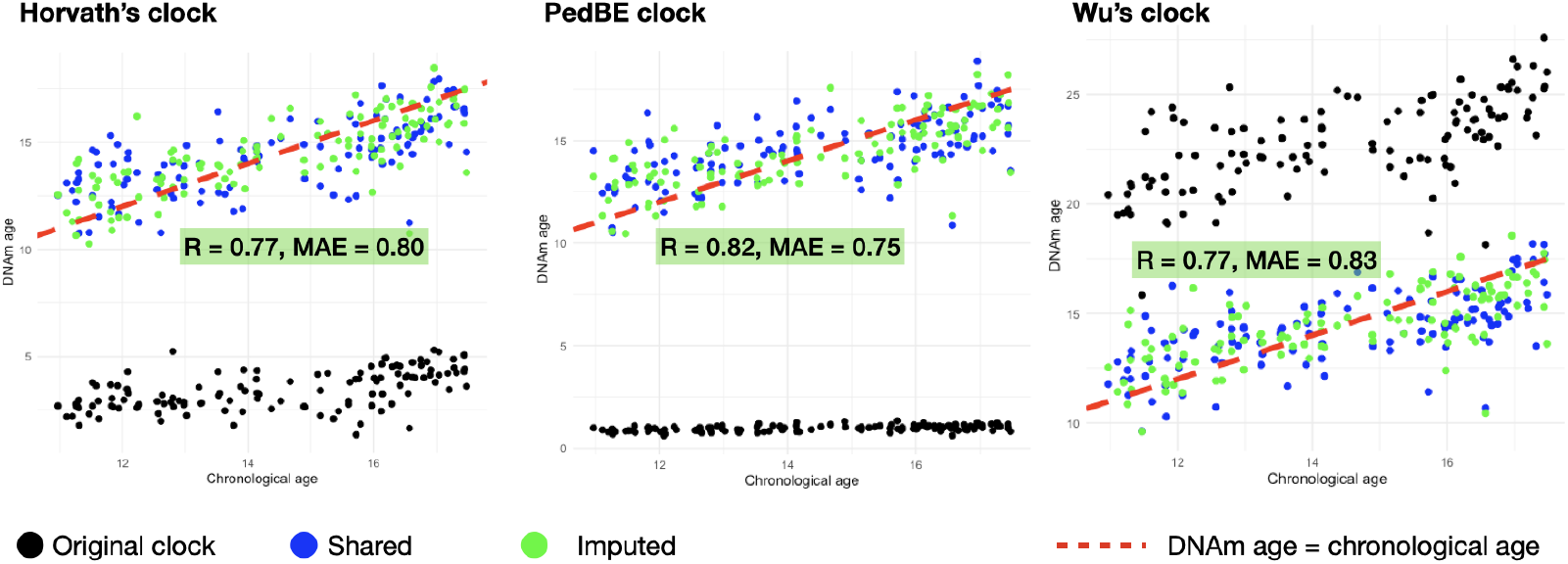
Plots for samples in test sets using the original clock, the transferred clocks with shared CpG sites, and the transferred clocks after feature alignment via imputation.

### 3.5 Transfer learning via deep neural networks

The idea of transfer learning in the DNN framework is different from that of the penalized linear regression model. Here, instead of further fine-tuning the coefficients of the profiled CpG sites, we assume that the selected CpGs and their weights in existing clocks are transferrable to our task. In other words, instead of tailoring 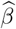 to our target dataset, we find the mapping from our input feature vector *z* ∈ ℝ^*q*^ to their latent feature representations through complex nonlinear functions. In summary, the neurons in the penultimate layer serve as a compact, high-level abstraction of the original input features, where each of them represents a learned component that contributes to the final age prediction. Unlike a single CpG site in traditional epigenetic clocks, each neuron in the penultimate layer may capture significant interactions among multiple CpG sites rather than reflecting the methylation level at one specific locus.

Since the sample size of our target dataset is limited, we first apply the sure independence screening technique (Fan and Lv, 2008) to reduce the dimension of the input vector by selecting the top 600 CpG sites. The input information is then processed through three hidden layers with 256 neurons in the first and 128 in the second layers, respectively. The number of neurons in the third layer (also the penultimate) equals the number of selected CpG sites in an existing clock; for example, it is 111 neurons if we transfer the Wu’s clock (Wu et al., 2019). All hidden layers employ the ReLU activation function to introduce nonlinearity into the model. The final layer is a single-unit dense layer with a linear activation function. We emphasize that the bias and weights connecting the penultimate layer and the final output layer are also fixed according to the publicly available coefficients in an existing clock. Similarly to the loss functions used to build existing clocks, the model uses the mean squared error loss function, and it is optimized with the Adam optimizer (Zhang, 2018).

Compared to the methods in Section 3.3 and Section 3.4, transfer learning via the proposed DNN outperforms linear models with shared or imputed CpG sites. Consider Wu’s clock, for example, the correlation coefficient improves up to 15.7% and the MAE decreases up to 32.6%. We expect further improvements if our target set has a larger sample size so that more CpG sites could be taken in the input vector, as reported in de Lima Camillo et al. (2022) that the expanded feature set helps improve performance.

## 4 Discussion

The past decade has witnessed the development of various epigenetic clocks to estimate the biological age of an individual based on cellular DNA methylation levels (Horvath, 2013; Hannum et al., 2013; Levine et al., 2018; Lu et al., 2019; Belsky et al., 2022). When it comes to scientific studies, researchers typically choose one or several of the most popular epigenetic clocks and apply them to their target cohorts. However, compared to the samples from the target cohort, these existing clocks can be trained with data sampled from a different underlying population, tissue, or cell type. Besides, the difference between the predicted age (also known as the DNAm age) and the true chronological age is referred to as epigenetic age acceleration, which has been viewed as an indicator of faster aging or a high risk of developing aging-related diseases (Jain et al., 2022; Tian et al., 2023; Cortez et al., 2024). However, if the estimated DNAm age differs markedly from the true chronological age, the clock is not useful, and the value of age acceleration may no longer be interpretable. In addition to the heterogeneity in the relationship between age and DNAm levels between source and target datasets, the CpG sites that are being profiled may be quite different due to the advancement of platforms. In particular, most existing clocks are trained with samples profiled by the older 27K or 450K array, while our target data is profiled by the more recent EPIC array. In other words, the number of CpG sites in our target data is significantly expanded on the 450K, and the feature dimension is much larger than that allowed by existing clocks.

To address the lack of generalizability of existing epigenetic clocks, we propose several methods for transfer learning within the framework of epigenetic clocks. These methods adapt existing epigenetic clocks trained from abundant source datasets to our target study. They do not require accessing the original raw data in the source domain, but only the profiled CpGs and their weights. To address cross-platform heterogeneity, we consider the imputation-based feature alignment method and the DNN-based method. Compared to the baseline method using only shared CpGs between 27K/450K and EPIC arrays, both methods show improved prediction performances.

Although our proposed transfer learning framework was mainly focused on first-generation clocks, we expect it to be generalizable to second-generation clocks to predict mortality risk and healthspan (Levine et al., 2018; Lu et al., 2019). In terms of statistical methodological developments, this generalization concerns transfer learning that goes beyond the high-dimensional linear regression models, for example, in Cox regression models and generalized linear models. Although existing work has considered transfer learning in these settings, it focuses on homogeneous transfer settings where the covariate sets are exactly the same between the source and target domains (Tian and Feng, 2023; Li et al., 2023). This may not be well aligned with the application of second-generation clocks, as the target study may only have partially overlapped covariate sets with the source datasets. For example, maybe some of the clinical biomarkers in the PhenoAge clock (Levine et al., 2018) have not been measured in the target study, or additional factors such as environmental exposures should be incorporated into the clock in the target study. Furthermore, the outcomes of interest can also differ between source and target studies, which requires further studies on how to ensure effective knowledge transfer. In summary, heterogeneity transfer learning might be a more prominent problem in second-generation clocks.

In addition to the underlying differences between populations with different demographic characteristics and environmental exposures, the gap between an existing clock and a target study can also arise from cell-type heterogeneity. To date, epigenetic clocks have been trained from bulk tissue samples, which are composed of many different cell types. For example, DNAm data sets in training Horvath’s clock and Wu’s clock, for example, are predominately extracted from whole blood or other cell types that display age-related changes similar to those observed in whole blood (Teschendorff and Horvath, 2025). Thus, existing epigenetic clocks should be viewed as a composite measure of two components, one reflecting the age-related change in cell-type composition of the tissue and another reflecting age-related DNAm changes of each cell type in the tissue. First, cell-type composition changes with age. For example, it is well known that the natural aging process is associated with a shift from lymphoid to myeloid cell production (Chinn et al., 2012). In addition, the rate of aging also differs between cell types. For example, hepatocytes in the liver of individuals with obesity could age faster than the surrounding liver stromal cells (Teschendorff and Horvath, 2025). Therefore, to better understand the rate of aging, it is of key scientific interest to disentangle cell-type convolutions. Developing cell-type-specific epigenetic clocks could better reveal the intrinsic aging rate induced by DNAm changes. One possible future direction is to explore the transferability between clocks trained with different tissue or cell types. In addition, it is also important to develop transfer learning methods for tailoring bulk tissue-based epigenetic clocks to a specific tissue or cell type.

## Data Availability

All data produced in the present study are available upon reasonable request to the authors.

## Notes

### Competing Interest Statement

The authors have declared no competing interest.

### Funding Statement

Research reported in this publication was supported by the National Institute On Aging of the National Institutes of Health under Award Number R21AG083364.

### Author Declarations

IRB of Rutgers University gave ethical approval for this work.

